# Combined virtual reality social cognition training and theta transcranial alternating current stimulation: A proof-of-concept study in healthy controls

**DOI:** 10.1101/2025.05.09.25327302

**Authors:** Kirsten Gainsford, Bernadette M. Fitzgibbon, Aron T. Hill, Paul B. Fitzgerald, Caroline T. Gurvich, Kate E. Hoy

## Abstract

Social cognition, particularly theory of mind (ToM) is important for understanding and engaging with the social environment. The continual development and improvement of these skills can be of broad benefit. However, current social cognitive training methods are time intensive and have limited ecological validity. Virtual reality (VR) combined with transcranial alternating current stimulation (tACS) may provide a more ecologically valid and efficient alternative. The current study investigated the effects of theta tACS to the right temporoparietal junction (rTPJ) on VR social cognition training in a group of 21 healthy adults. Outcome measures were behavioural (attribution of intentions ToM task) and neurophysiological (spectral power and event related potentials). Participants completed two identical lab sessions each. One session included VR training concurrent with active theta tACS and the other, with sham theta tACS. Participants completed resting state electroencephalography (EEG) and ToM tasks with concurrent EEG pre and post VR-tACS in each session. tACS condition was randomised and all assessments were double-blinded. VR and active tACS, but not VR and sham tACS, improved ToM task accuracy. ToM task response times improved pre versus post VR-tACS regardless of tACS condition (active vs sham). Resting state theta power increased significantly across the cortex post VR-tACS regardless of tACS condition. This study provides the first evidence for the feasibility of a combined VR-tACS protocol for social cognition. Future research in larger samples, and with multiple sessions, in both healthy and clinical populations are recommended.

## 1. Introduction

Social cognition, which includes theory of mind (ToM), refers to a set of skills crucial for understanding, predicting and engaging with the world and people around us (Arioli, Crespi, & Canessa, 2018; Green, Horan, & Lee, 2019). ToM is the ability to take the perspectives of others and predict and interpret their intentions and behaviours as well as infer meaning from them (Green et al., 2019) Social cognition develops from infancy and continues to evolve throughout the lifespan (Arioli et al., 2018; Roheger, Hranovska, Martin, & Meinzer, 2022). The development of social cognitive skills is impacted by highly dynamic factors such as environment, upbringing, socioeconomic status, stress and relationships (Charness, List, Rustichini, Samek, & Van De Ven, 2019; Hughes et al., 2005; Nolte et al., 2013; Rakoczy, 2022; Roheger et al., 2022). Because of this, there is considerable variation in social cognitive abilities in the healthy population (Roheger et al., 2022).

Social cognitive training can assist in improving many aspects of day-to-day functioning and development such as improving emotional well-being and building strong relationships (Charness et al., 2019; Funghi et al., 2024; Schoeneman Patel et al., 2022). Programs to enhance social cognition, including in the healthy population, generally involve engagement in computer– and/or group-based therapies to target specific or broad social cognitive skills (Roheger et al., 2022). While evidence shows these programs can lead to some improvements in social cognitive abilities, they are often time intensive and resource-heavy (Roheger et al., 2022). These interventions are also generally carried out at schools, aged care facilities, and clinical settings with the intention that they will generalise to the environments that trained skills are relevant to (Gainsford, Fitzgibbon, Fitzgerald, & Hoy, 2020; Roheger et al., 2022; Yeo, Yoon, Lee, Kurtz, & Choi, 2022). However, spontaneous generalisation from these training settings to real-world applications rarely occurs (Cavallini et al., 2015; Roheger et al., 2022). Virtual reality (VR) is being increasingly used for training social cognition with promising results (Pérez-Ferrara et al., 2024). It is ecologically valid, allowing people to be immersed in environments created to reflect the ones learned skills will be used in, while also allowing control of these environments to personalise training and skills development (Parsons, Gaggioli, & Riva, 2017). Additionally, it has been shown to be as (or more) effective for training social cognition skills in fewer sessions than traditional methods (Gainsford et al., 2020; Pérez-Ferrara et al., 2024). As the evidence for VR-based training efficacy grows, researchers have been integrating it with other technologies such as non-invasive brain stimulation (NIBS) to further enhance effects, when treating conditions such as cerebral palsy and stroke (Cassani, Novak, Falk, & Oliveira, 2020).

NIBS refers to a group of neuromodulation techniques that can be used to prime brain activity prior to VR intervention or used concurrently with VR (Cassani et al., 2020). Overall VR-NIBS literature suggests the combined approach may have an advantage over and above either VR training or NIBS alone (Cassani et al., 2020). Transcranial alternating current stimulation (tACS) is a type of NIBS that introduces an alternating electrical current at a chosen frequency to entrain brain activity (Antal & Paulus, 2013). The ability to entrain brain activity to specific frequencies is of particular interest in social cognitive training as theta brain activity (4-8Hz) has been associated with engagement in ToM processes (Christian, Kapetaniou, & Soutschek, 2023; Wang, Callaghan, Gooding-Williams, McAllister, & Kessler, 2016). With theta band synchronisation increasing across frontal (i.e., lateral prefrontal cortex, anterior cingulate cortex) and parietal (right temporoparietal junction [rTPJ]) social cognitive networks when engaging in ToM (Seymour, Wang, Rippon, & Kessler, 2018; Uhlhaas, Haenschel, Nikolic, & Singer, 2008). Modulation of theta activity using tACS has been shown to improve ToM (Christian et al., 2023). Christian et al. (2023) administered a single session of theta, beta or sham tACS to the rTPJ (a hub of ToM processing) of healthy participants during a ToM perspective taking task, finding that following theta tACS participants were more likely to make decisions during the task that took into consideration the wellbeing of others.

To date, no studies have combined VR and tACS to improve social cognition. Therefore, the current study investigated, for the first time, the effects of combined VR social cognitive training and theta tACS to the rTPJ in a healthy sample, looking at both behavioural and neurophysiological outcome measures. We hypothesised that there would be a significantly greater improvement in ToM task accuracy and response time after VR social cognition training and active theta tACS (VR+tACS) compared to VR and sham tACS (VR+Sham). Additionally, we hypothesised that VR+ tACS, compared to VR+Sham, would result in increased theta power at rest aswell as increased mean amplitude of TP450 and late positive component (LPC) event-related potentials (ERP) at the rTPJ while completing ToM tasks. These ERPs are the neural responses commonly seen when the brain engages in ToM processes (Libsack et al., 2022; Vistoli et al., 2015).

## 2. Methods

### 2.1 Participants

Twenty-one healthy adult participants (12F, 9M, M = 33.62 years) were recruited to the study. Participants were excluded if they had a history of seizures or epilepsy, neurological disorders, significant traumatic brain injury or if they had substance use disorders or other unstable medical or psychiatric conditions. The study took place at the Epworth Centre for Innovation in Mental Health (ECIMH) between 2021 and 2022, and then the Monash Alfred Psychiatry Research Centre (MAPrc) between 2022 and 2023. This was due to the closure of ECIMH in 2022. Additional details in supplementary materials.

Participants were recruited via social media (e.g., Facebook, X), recruitment websites (e.g., HealthMatch) and flyers posted at universities and community centres. All participants provided written informed consent. Ethics approval was obtained from the Monash Health Human Research Ethics Committee and governance obtained through Monash University, Alfred Health and Epworth HealthCare.

### 2.2 Design

This was a double-blind, randomised, controlled, within-subjects study. Participants attended three sessions in total. At the baseline visit (i.e. Session 1) participants completed a cognitive and social cognitive task battery. Data from the baseline visit, is not the focus of the current paper and has been published previously (Gainsford et al., 2025). Sessions two and three are outlined in **Figure 1**. Sessions were conducted at least 72 hours apart. Participants were randomly allocated to active or sham tACS for Session 2 and the opposite protocol was applied in Session 3. Participants wore an EEG cap embedded with tACS electrodes at all times during Sessions 2 and 3. This stayed on the participant’s head before, during and after VR-tACS. EEG cords were stowed in a backpack while the participant completed VR-tACS. Detailed tACS blinding and randomisation information is outlined in **Section 2.6** below.

**Figure 1:**
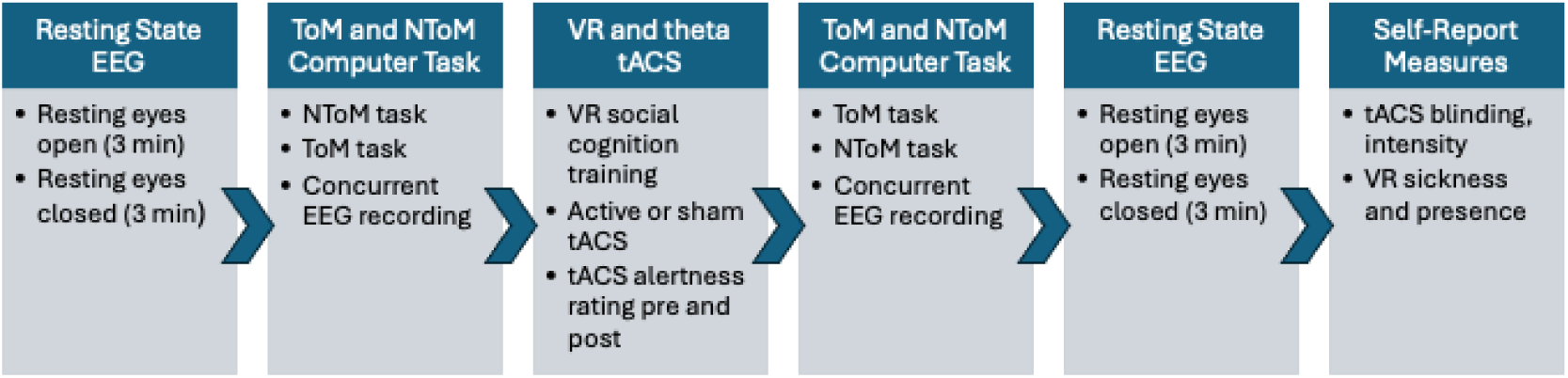
Study processes for Sessions 2 and 3. Participants first had EEG recorded while resting with eyes open, then eyes closed. They completed the non-theory of mind (NToM) task then theory of mind (ToM) task. Then tACS impedance was conducted, VR headset put on and VR-tACS completed. Once finished, participants repeated the computer tasks with ToM first, then resting EEG measures. Participants then completed self-report questionnaires regarding their experience of VR and tACS.

### 2.3 EEG

EEG data were collected using a 50-channel montage (see supplementary materials) from the 10-05 system (ActiCap, Brain Products, Gilching, Germany) via Brain Vision Recorder software (Brain Products, Gilching, Germany) with Ag/AgCl electrodes connected to a BrainAmp amplifier (Brain Products, Gilching, Germany). The EEG montage was customised to accommodate embedded tACS electrodes and the VR headset. The following electrodes were used: AF3, AFz (ground), AF4, F7, F5, F3, F1, Fz, F2, F4, F6, F8, FC5, FC3, FC1, FCz, FC2, FC4, FC6, T7, C5, FCC6h, CCP5h, C3, C1, Cz, C2, C4, FCC6h, CCP6h, T8, TP7, CP5, CP3, CP1, CPz (reference), CP2, CP4, CP6, TP8, P7, P5, P3, P1, Pz, P6, P8, PO3, POz, PO4, O1, Oz and O2. Impedances were kept below 10 kΩ and data were collected at a sampling rate of 1000Hz with a 0.01Hz high-pass filter and low-pass filter of 200Hz.

The EEG cap stayed on the participant’s head throughout the session. While completing the VR-tACS protocol, the electrode cords were unplugged and stowed in a bag on the participant’s back. Immediately after VR-tACS was completed, the VR device was removed, the person was either moved back into the EEG lab or lab equipment moved back into position, EEG was plugged in, impedance checked, and the ToM computer task was started. Participants started the ToM task within approximately five minutes of completing VR-tACS. ToM task version presentation was counterbalanced.

#### 2.3.1 EEG Resting State

In each session, three minutes of resting eyes open and three minutes eyes closed was recorded prior to concurrent VR and tACS as well as after. Participants were seated approximately 1m from the computer screen and were instructed to sit as still as possible during recordings. They were instructed to look at a fixation cross in the centre of a computer screen for resting eyes open and told to relax with eyes closed for resting eyes closed.

### 2.4 Behavioural Task

Participants completed the attribution of intentions ToM task prior to, as well as after VR-tACS in each session. This task is commonly used to investigate mental state reasoning (Eddy, 2019). EEG was simultaneously recorded each time the task was completed. The task was built on Inquist 6 (Millisecond, Seattle WA, USA, 2020) based on the one used by Vistoli et al. (2015) using the same stimuli. However, due to having multiple sessions, stimuli were separated into multiple task versions.

There are 82 comic strips in total, each with four pictures including a congruent and incongruent ending. There are three types of comics, intention attribution requiring use of ToM to interpret (i.e., ToM task), physical causality with objects (PCOb) and physical causality with characters (PCCh), the latter two not requiring the use of ToM to interpret. PCOb and PCCh stimuli were combined to form the non-theory of mind task (NToM).

There were two ToM task versions (ToM A, ToM B) and two NToM task versions (NToM A, NToM B). Each ToM task contained 24 trials (12 congruent endings, 12 incongruent endings) and each NToM task contained 48 trials (24 congruent endings, 24 incongruent endings). The remaining ten comics formed two shorter practice tests. Each task version (A or B) contained either the congruent or incongruent comic ending. Comics were randomly allocated into task versions using an online number generator and were not repeated within task versions. Order of comic presentation was randomised within each task, each time it was presented. Task order was counterbalanced. Participants saw each version of each task once per session.

To complete the tasks, participants sat in a chair approximately one metre from the computer screen. For the ToM task, they were given instructions to consider whether the fourth picture in each comic “makes sense for the person’s goals”. For the NToM task, participants were instructed to consider if the fourth picture “makes sense as the ending of the story”. Participants pressed a green button on a button box if the fourth picture made sense and a red button if it did not. The time sequence of the comics is shown in **Figure 2**. The fourth picture was presented until the participant responded or for 4000ms, whichever came first. The fourth picture of each comic had a red square around it to assist participants in knowing when to respond (**Figure 2**). Number of correct responses and response times were recorded.

**Figure 2:**
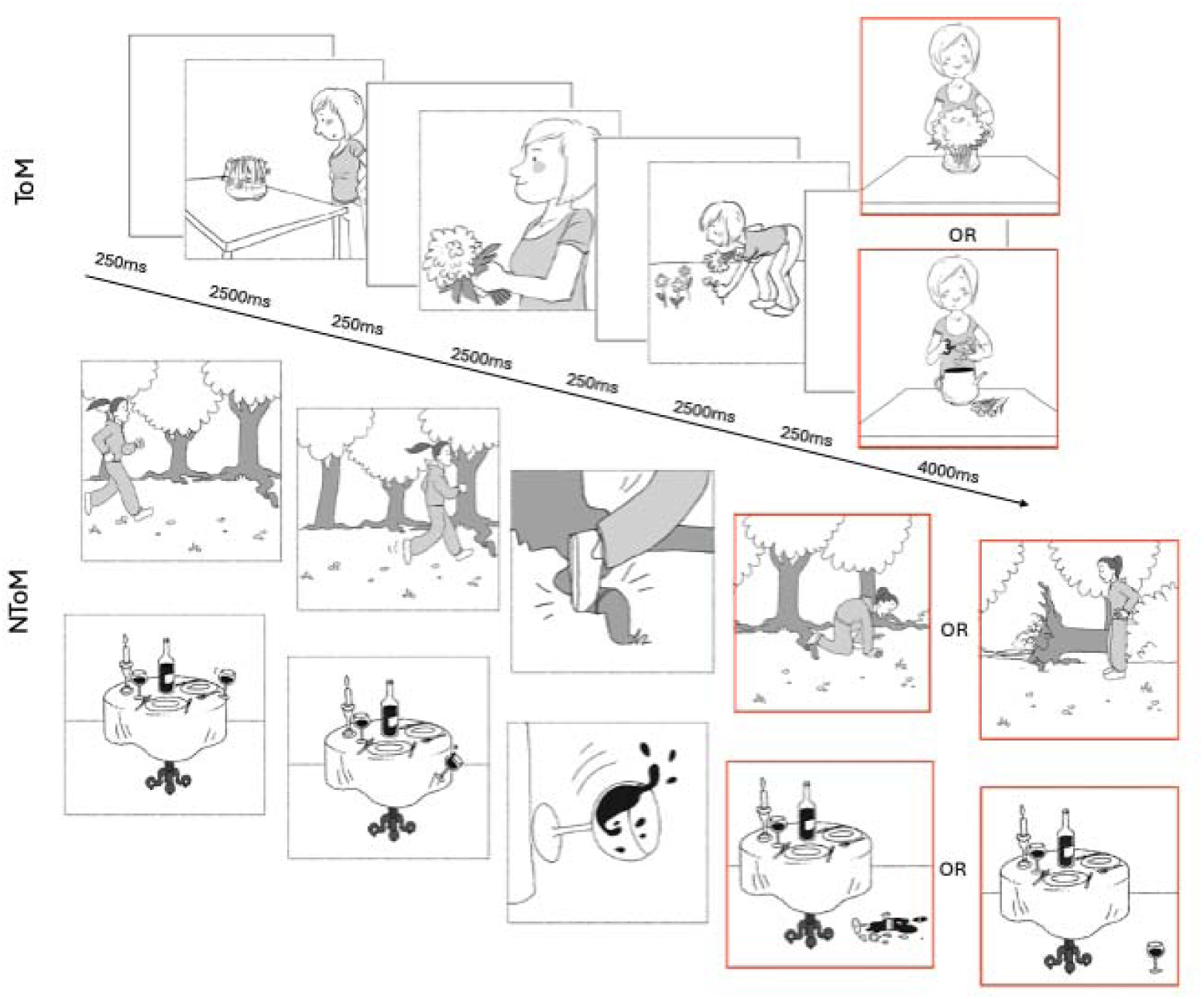
Attribution of intentions ToM task time course. The first is a ToM example with congruent or incongruent ending options and NToM examples (PCCh and PCOb) with alternate endings below (Reproduced from Gainsford et al., 2025)

### 2.5 VR

Participants wore a Meta Quest 2 head-mounted display (HMD; Meta, 2020) with hand-held controllers. At both research sites, lab set up ensured participants were able to move freely and safely within the space while completing the VR task. Participants navigated through two virtual environments (shopping mall and restaurant, **Figure 3**) in a custom-built VR program (SoCog, SensiLab, Monash University, Melbourne, Australia). Participants engaged with virtual avatars to interpret their actions or emotions and decided how to respond from a list of options provided. The program gave feedback on whether the responses were correct, aiding learning. Participants completed the VR tasks once during active and sham tACS sessions respectively (twice in total).

**Figure 3:**
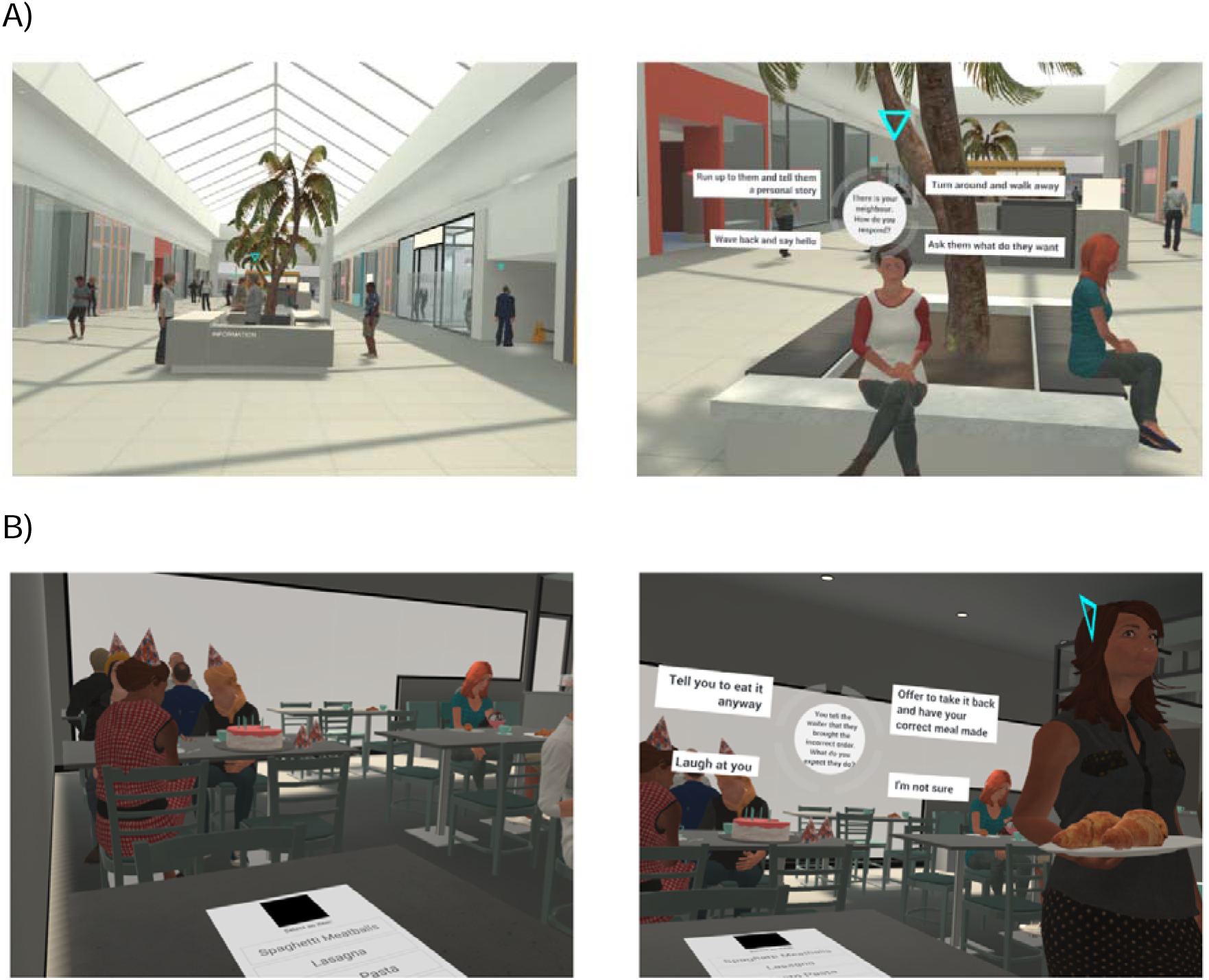
Examples of virtual environments participants encountered in VR social cognition training. A) Participants explored a shopping mall while encountering tasks such as the one on the right where they had to determine how they would respond in particular social situations based on social conventions. B) Participants had a meal at a café and were required to manage difficult situations such as determining how to respond when the waiter brings the incorrect food order.

### 2.6 tACS

Active tACS included 15 minutes of 5Hz theta frequency stimulation with a 30 second ramp up in intensity at the start of stimulation and a 30 second ramp down at the end (16 min total). The sham condition comprised of a 30 second ramp up followed by an immediate 30 second ramp down. This is a commonly used sham condition to provide the sensation of receiving tACS without the neurophysiological effects. The SimNIBS software (Version 3.2) was used to conduct E-field current modelling (Saturnino, Madsen, & Thielscher, 2020). A montage was developed using a combination of trial-and-error and MNI co-ordinates to achieve maximal current flow over the cortical regions corresponding to the rTPJ (MNI coordinates: 50, –53, 35, **Figure 4**). To select stimulation intensity, a small pilot of three intensities was conducted using the selected montage (C4, C6, P2, P4) prior to commencing the study. The total current was 1.75mA, tACS was administered using the StarStim^®^ wireless hybrid EEG/transcranial current stimulation 8 channel system (Neuroelectrics, Barcelona, Spain), and NIC2 program (Neuroelectrics, Barcelona, Spain). Pi stim Ag/AgCl (3.14cm^2^) electrodes were filled with a saline gel and tACS impedance was kept below 10kΩ.

**Figure 4:**
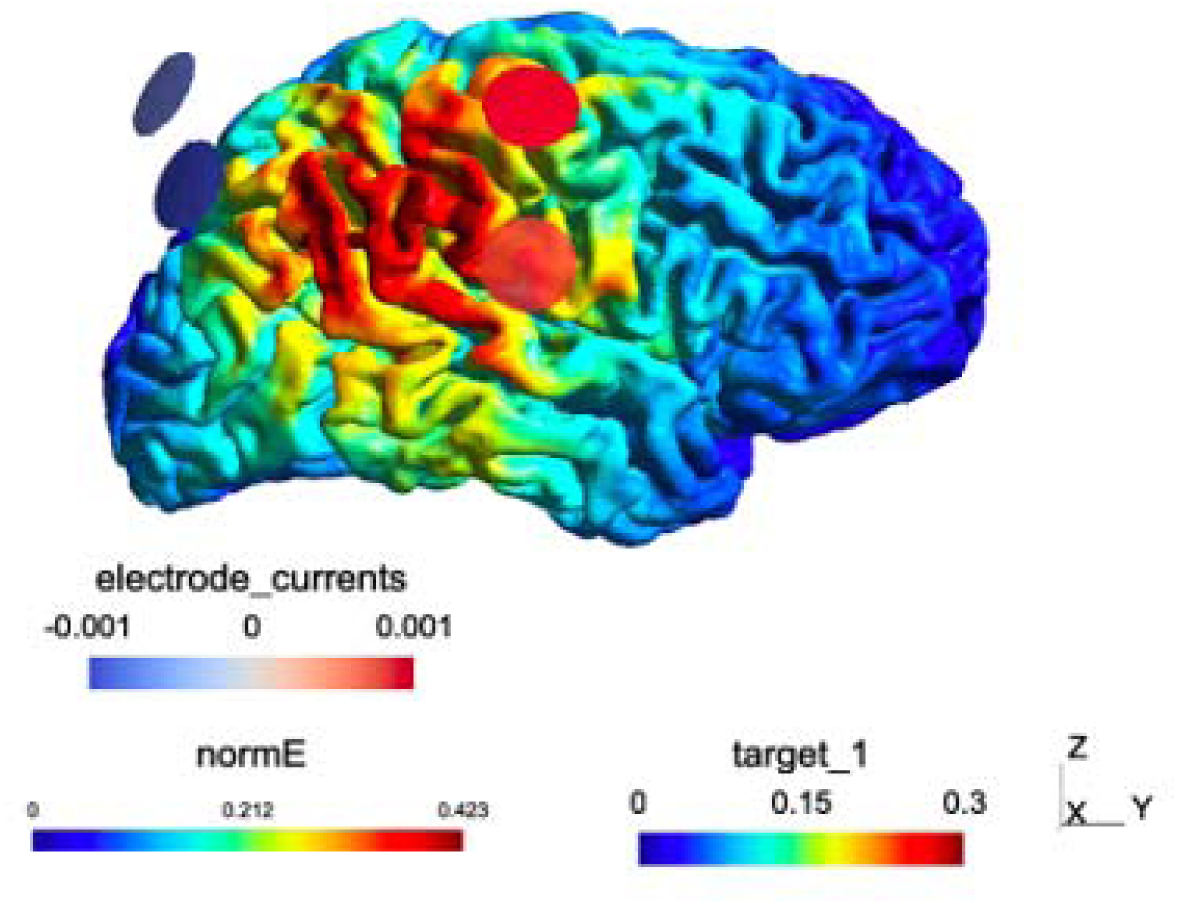
Current modelling conducted with SimNIBS to determine electrode placement for theta tACS. Electrodes were embedded in the EEG cap at C4, C6, P2 and P4. Red circles represent stimulation electrodes (C4, C6) and blue circles represent return electrodes (P2, P4).

The active and sham protocols were added to the NIC2 program and an unblinded team member not involved in data collection or analysis, assigned a code to each protocol (A or B). The double-blind mode was turned on using a password only accessible to the unblinded researcher. Participants were randomly allocated to active or sham tACS for Session 2 and the opposite protocol was applied in Session 3. Stimulation protocols were randomly allocated using Microsoft Excel, randomly assigning letters A or B against a number from 1-26. Each participant received one active and one sham session of tACS. Participants were set up in the virtual environment and began the VR tasks once tACS ramp up period had ended (30 sec). Participants completed the VR task while receiving tACS. If the VR task was completed prior to tACS finished, participants were sat down in a calm virtual environment until the tACS ended.

### 2.7 tACS Blinding and Intensity

Participants were asked in each experimental session to guess whether they were receiving active or sham stimulation and to rate their confidence on their guess ranging from 0 (“complete guess”) to 10 (“absolutely sure”). After stimulation, participants also rated intensity of stimulation (0, “no sensation” to 10 “extremely strong”) and provided feedback on any other sensations or side effects they felt during stimulation.

### 2.8 VR Sickness and Presence

The VR sickness questionnaire (VRSQ; Kim, Park, Choi, & Choe, 2018) was completed by participants after VR-tACS in each experimental session. Participants rated the degree to which they experienced side effects during the VR task on a scale of 0 (“not at all”) to 3 (“very”). Oculomotor (general discomfort, fatigue, eyestrain, difficulty focusing) and disorientation (headache, fullness of head, blurred vision, dizziness, vertigo) sub-scores were calculated, and total scores calculated from these. The presence questionnaire (PQ; Witmer & Singer, 1998) was also completed by participants at the end of each session to assess the level of immersion they experienced in the VR. Each question had a Likert scale of 1-7 (seven being the highest). Sub-scores were calculated by adding up total scores for items relating to realism, possibility to act, quality of interface (reverse-scored), possibility to examine, self-evaluation of performance, and sounds.

### 2.9 Analysis

#### 2.9.1 Task Response Accuracy and Response Time

To determine response accuracy, the percentage of correct responses was calculated and averaged for each participant. Mean response times were calculated by extracting the time at which participants responded to the fourth picture in each comic and averaged. Trials were excluded from analysis where participants responded on the button box before the presentation of the fourth image of the comic (i.e., response error). Using JASP (Version 0.17.3, JASP Team, Amsterdam, Netherlands 2023), pairwise t-tests were conducted for pre versus post active and sham VR-tACS accuracy and response time. To examine the effects between stimulation conditions, difference scores were calculated by subtracting pre-VR-tACS task scores and response times from post-VR-tACS task scores and response times for active and sham, and the difference scores were statistically compared using paired samples t-tests.

#### 2.9.2 EEG Analysis

All EEG data were pre-processed using RELAX v1.1.5 (Bailey, Biabani, et al., 2023; Bailey, Hill, et al., 2023), a fully automated EEG pre-processing pipeline, in MATLAB (2024a, Mathworks, 2024) with the EEGLAB toolbox (Delorme & Makeig, 2004). As part of the RELAX pipeline, the data were zero-phase Butterworth bandpass filtered. The high– and low-pass filters for the resting state data was 0.5-80Hz and 0.25-80Hz for ERP analysis. The lower high-pass filter for ERPs is used to avoid any potential distortion of the ERPs from more aggressive filtering (Bailey, Hill, et al., 2023). A 50Hz notch filter was then applied to remove line noise. Bad electrodes were initially removed by the PREP pipeline (Bigdely-Shamlo, Mullen, Kothe, Su, & Robbins, 2015). Then multi-channel Wiener filtering applied to clean muscle activity, blinks and horizontal eye movements and were re-referenced (Somers, Francart, & Bertrand, 2018). Independent component analysis (ICA; FastICA-symm method) was then performed to identify remaining artifacts using the ICLabel classifier (Pion-Tonachini, Kreutz-Delgado, & Makeig, 2019) and then removed using wavelet enhanced ICA (wICA; Bailey et al., 2024). Any rejected electrodes were then re-interpolated using spherical interpolation.

For resting state analysis, the cleaned data were separated into 3 second epochs (–1.5 to 1.5 sec) with no overlap (see additional information in supplementary materials). For ERP analysis, epochs were created around the third image of each comic as this was the point of the story where ToM neural processes would be activated (Vistoli et al., 2015). Epoch lengths for ERP analysis were 1.3 seconds (–300ms to 1000ms, additional information in supplementary materials). FieldTrip (Oostenveld, Fries, Maris, & Schoffelen, 2011) was used to average each participants’ epochs from pre– and post-tACS for resting data and ERP data to create grand average files used to conduct analyses.

##### 2.9.2.1 Resting State EEG Cluster Analysis

Pre-processed and epoched data were transformed into the frequency domain using a fast Fourier transform (FFT) of 1-45Hz at 0.5Hz intervals to calculate spectral power. Grand average files were calculated for each condition (resting eyes open and closed, pre and post, active and sham). Resting state cluster-based permutation analyses were conducted using a whole-brain data driven approach, to broadly identify areas of cortical activation at rest and consisted of 5000 permutations. To determine change in theta power (4-8Hz) at rest, within-subjects cluster analyses were conducted for pre vs post active and sham using customised MATLAB scripts. To determine effect of stimulation on resting state theta power post VR+tACS and post VR+Sham, resting data were statistically compared using two-tailed within-subjects (dependent samples) t-tests (α = 0.05, two-tailed).

##### 2.9.2.2 Resting State EEG ROI Analysis

A group of electrodes representing the rTPJ (CP4; CP6; TP8; P6; P8) were selected to analyse theta band activity (4-8Hz) at rest. Mean amplitudes at these electrodes were extracted for each participant and inputted into JASP. To determine change in resting state theta power post VR-tACS compared to pre within active and sham sessions, data were statistically compared using paired samples t-tests. To compare active and sham conditions, post-VR and tACS, mean amplitudes were subtracted from pre-to create difference scores for active and sham sessions. The data was then compared in JASP using paired samples t-tests.

##### 2.9.2.3 Event-Related Potential Analysis

Two ERPs were investigated that relate to ToM processes. First, we examined the temporoparietal 450 (TP450), peaking at approximately 450ms, with the analysis timepoint of interest classified as 325ms to 525ms. To conduct ROI analyses, we investigated changes in peak amplitude at the CP6 electrode (representing the rTPJ) and a broader group of electrodes around the rTPJ (CP4, CP6, TP8, P6, P8). We examined the late positive component (LPC) occurring centro-parietally between 300ms and 700ms. Based on the literature, changes in peak amplitude were analysed at Pz (parietal midline) alone and Pz and Cz together (centro-parietal midline; taken as an average of the signal across the two electrodes). To determine change in theta power post-VR-tACS compared to pre-within active and sham sessions, ROI analysis was conducted by extracting the mean amplitude across the aforementioned electrodes and statistically comparing them in a series of paired-samples t-tests using JASP. To compare active and sham conditions, post-VR and tACS, mean amplitudes were subtracted from pre-to create difference scores for active and sham sessions prior to conducting paired samples t-tests. Cluster-based analyses were also conducted as an additional ‘whole brain’ data-driven approach to explore which cortical regions activated during task performance. Two-tailed dependent samples t-tests were conducted to identify significant electrode clusters pre-versus post-VR-tACS within each session and post-versus post-VR+tACS and VR+Sham (α = 0.05, two-tailed; Maris & Oostenveld, 2007).

#### 2.9.3 Correlations

Any significant electrode clusters from resting state cluster-based permutation analyses were extracted using MATLAB and FieldTrip. Mean amplitude of theta power across these electrodes were calculated for each participant and each condition (resting eyes open and resting eyes closed, pre and post, active and sham). Difference scores were calculated using the extracted mean amplitudes by subtracting pre-from post-stimulation amplitudes for active and sham conditions. This provided scores indicating change in amplitude within sessions for active and sham. Pearson correlations were then performed comparing difference scores for resting eyes closed and resting eyes open active and sham with difference scores for ToM and NToM task accuracy and response times for active vs sham.

#### 2.9.4 tACS Intensity and Blinding

Active and sham blinding assessments were compared using chi square goodness of fit analyses. tACS stimulation intensity for active vs sham conditions were compared using a paired samples t-test. The researcher remained blinded until all data analysis was completed.

#### 2.9.5 VR sickness and immersion

VRSQ and PQ sub-scores and total scores were compared between active and sham conditions using paired-samples t-tests.

Assumptions of normality and variance were tested for all analyses and where these assumptions were violated, non-parametric tests were used and reported in the results section below. Note, Bayes Factor analyses (BF_10_) were conducted on all non-significant findings.

## 3. Results

### 3.1 Behavioural Results

#### 3.1.1 Task Response Time and Accuracy

##### 3.1.1.1 Theory of Mind task

In the within-session analysis, ToM task response accuracy significantly improved post-active VR-tACS compared to pre-active VR-tACS (*t*(20) = –3.055, *p* = 0.006) but did not significantly improve after sham VR-tACS compared to pre-(*t*(20) = –1.675, *p* = 0.110, BF_10_ = 0.751; see **Table 1** and **Figure 5**). Participants’ response times on the ToM task improved significantly post-active (*t*(20) = 7.484, *p* = <0.001) and sham VR-tACS (*t*(20) = 6.242, *p* = <0.001) compared to pre-. On the between session comparisons (i.e., active vs sham sessions), using difference scores between active and sham, there was no significant difference on response accuracy (*t*(20) = –1.255, *p* = 0.224, BF_10_ = 0.453) or response time (*t*(20) = 0.636, *p* = 0.532, BF_10_ = 0.273) between stimulation conditions.

**Figure 5:**
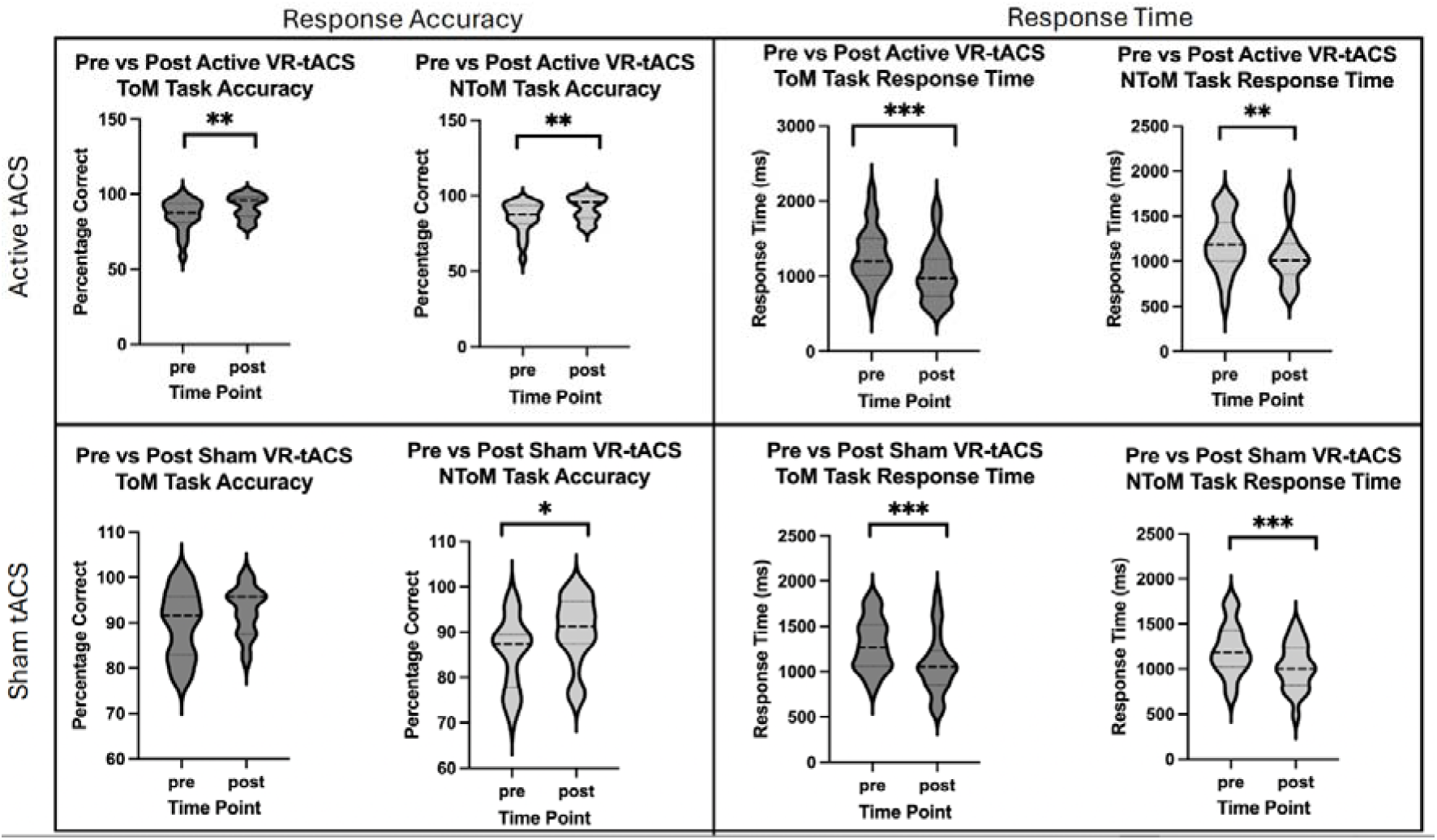
Comparison of response time and accuracy in ToM and NToM tasks pre vs post active and sham tACS combined with VR social cognition training. *p = < 0.05, **p = < 0.01, ***p = < 0.001

**Table 1:** ToM task response time and accuracy descriptive statistics.

|  | Accuracy |  |  | Response Time |  |  |
| --- | --- | --- | --- | --- | --- | --- |
|  | Pre | Post | Difference | Pre | Post | Difference |
|  | M(SD) | M(SD) | MD<sup>a</sup>(SD)<sup>b</sup> | M(SD) | M(SD) | MD<sup>a</sup>(SD)<sup>b</sup> |
| Active | 85.299<br>(10.408) | 91.589<br>(7.769) | 6.290<br>(9.436) | 1285.782<br>(365.833) | 1038.888<br>(365.00) | -246.894<br>(151.183) |
| Sham | 89.641<br>(7.374) | 92.785<br>(5.277) | 3.144<br>(8.604) | 1298.043<br>(285.805) | 1076.674<br>(328.819) | -221.369<br>(162.515) |

<sup>a</sup>mean difference, <sup>b</sup>standard deviation of mean difference

##### 3.1.1.2 Non-Theory of Mind Task

In the within-session analysis, NToM task response accuracy significantly improved post-active (*t*(20) = –3.180, *p* = 0.005) and sham (*t*(20) = –2.252, *p* = 0.036) VR-tACS compared to pre-. NToM task response time significantly improved post-active (*t*(20) = 3.341, *p* = 0.003) and sham (*t*(20) = 5.582, *p* = <0.001) VR-tACS compared to pre-(see **Table 2** and **Figure 5**). On the between session comparisons (i.e., active vs sham sessions), using difference scores between active and sham, there was no significant difference on response accuracy (Wilcoxon signed rank test; *z* = –0.523, *p* = 0.616, BF_10_ = 0.350) or response time (Wilcoxon signed rank test; *z* = –0.921, *p* = 0.374, BF_10_ = 0.563) between stimulation conditions.

**Table 2:**
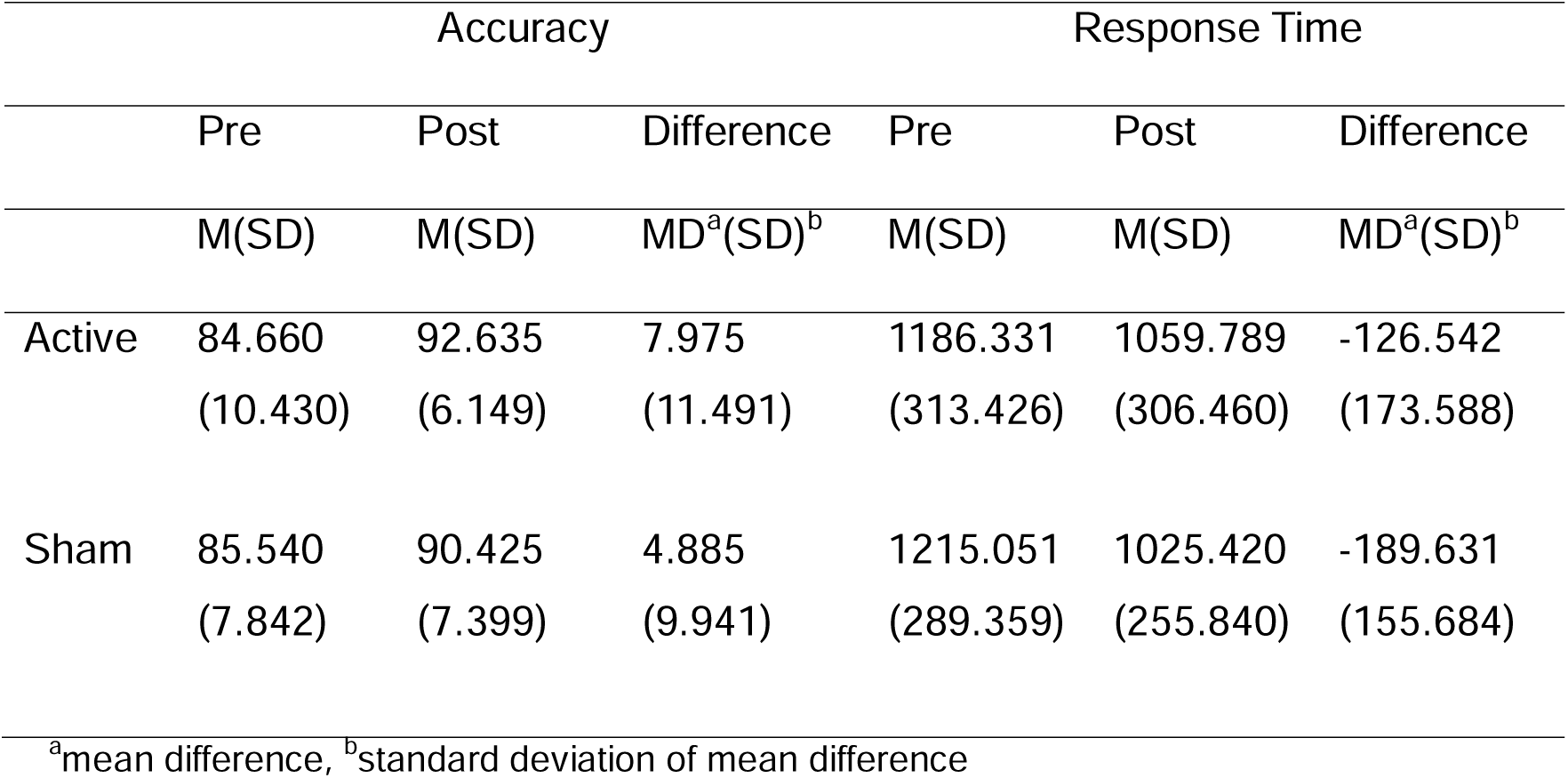
NToM task response time and accuracy descriptive statistics.

|  | Accuracy |  |  | Response Time |  |  |
| --- | --- | --- | --- | --- | --- | --- |
|  | Pre | Post | Difference | Pre | Post | Difference |
|  | M(SD) | M(SD) | MD<sup>a</sup>(SD)<sup>b</sup> | M(SD) | M(SD) | MD<sup>a</sup>(SD)<sup>b</sup> |
| Active | 84.660<br>(10.430) | 92.635<br>(6.149) | 7.975<br>(11.491) | 1186.331<br>(313.426) | 1059.789<br>(306.460) | -126.542<br>(173.588) |
| Sham | 85.540<br>(7.842) | 90.425<br>(7.399) | 4.885<br>(9.941) | 1215.051<br>(289.359) | 1025.420<br>(255.840) | -189.631<br>(155.684) |

<sup>a</sup>mean difference, <sup>b</sup>standard deviation of mean difference

### 3.2 EEG Analysis

#### 3.2.1 Resting State EEG Theta Power Cluster Analyses

##### 3.2.1.1 Resting Eyes Open

There was a significant increase in theta power post-active VR-tACS across all electrodes (p = 0.0008) as well as post-sham VR-tACS (p = 0.0002, **Figure 6A, 6B**). There was no significant change in theta power when comparing stimulation conditions (active vs sham, see supplementary materials).

**Figure 6:**
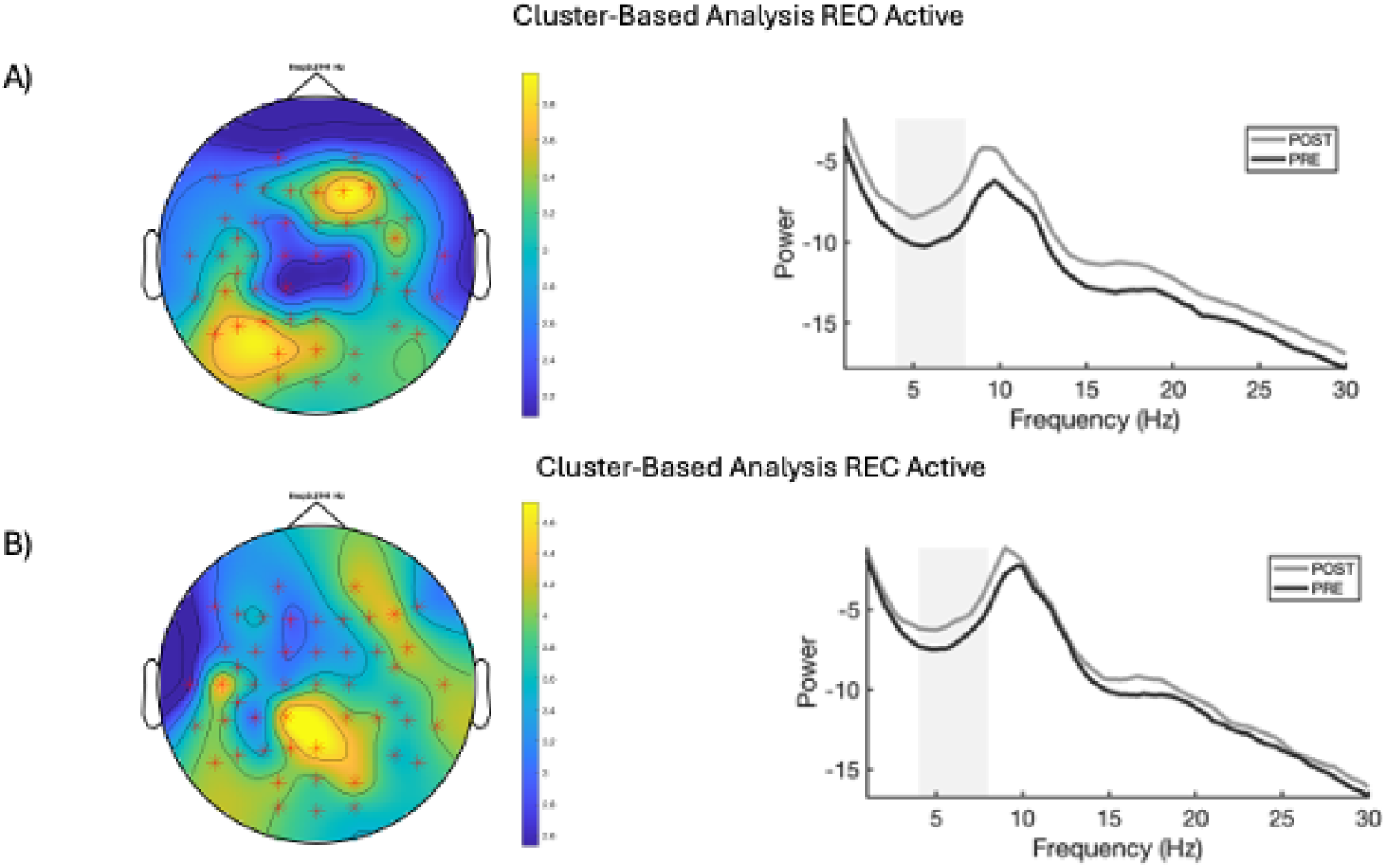
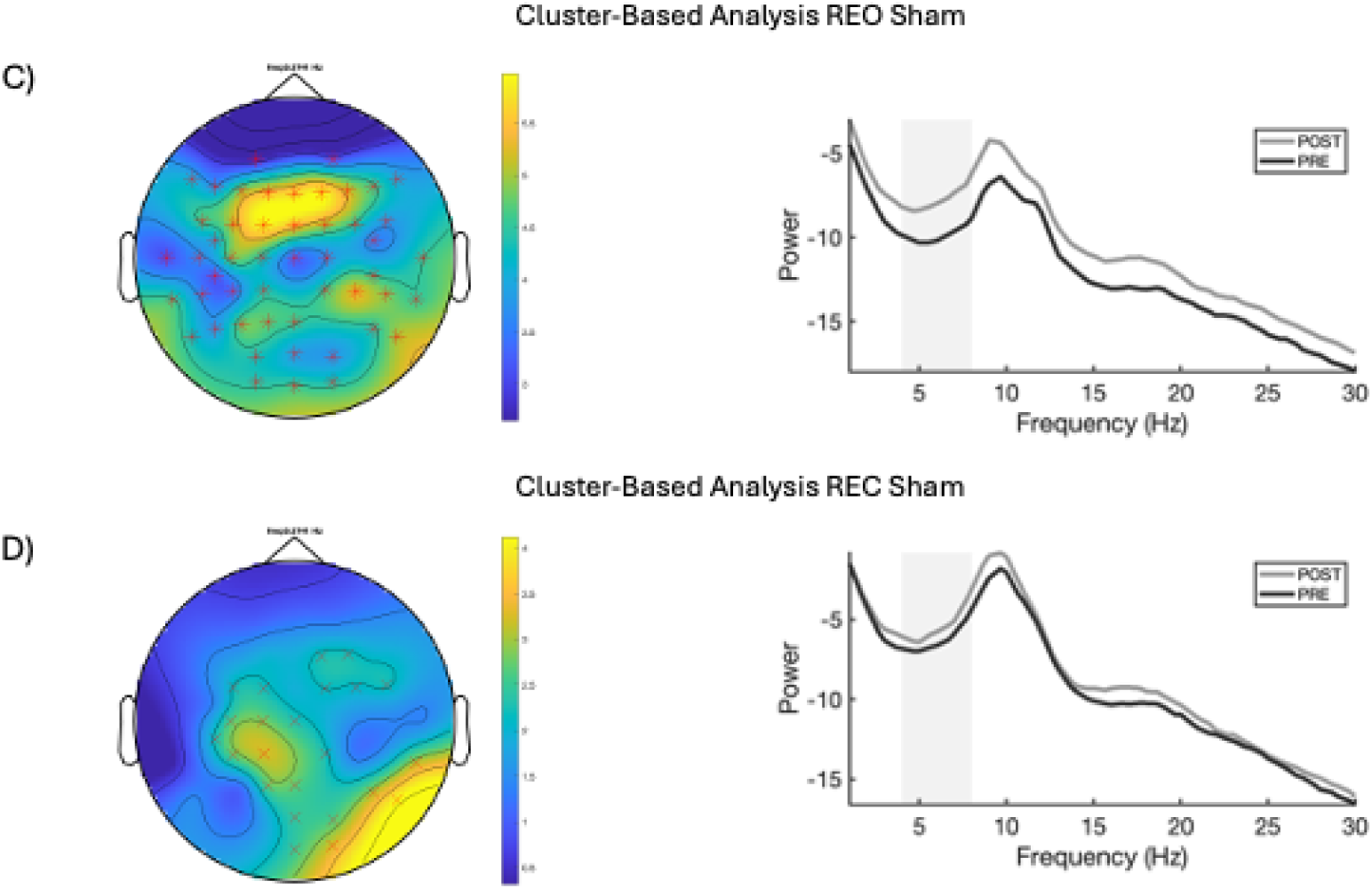
Cluster-based resting state EEG analysis comparisons. Figure 6A) Cluster analysis for pre-post VR+tACS, resting with eyes open. Figure 6B) Cluster analysis for pre-post VR+tACS, resting with eyes closed. Figure 6C) Cluster analysis for pre-post sham VR+Sham, resting with eyes open. Figure 6D) Cluster analysis for pre-post VR+Sham, resting with eyes closed. The spectral power plots provide an additional representation of the difference in brain activity at the significant clusters post-compared to pre-VR and tACS. The grey band on the spectral power plots highlight the theta activity band (4-8Hz). Significance levels on topoplots: ^x^p = < 0.05, * p = < 0.01

##### 3.2.1.2 Resting Eyes Closed

There was a significant increase in theta power post-active VR-tACS across all electrodes (p = 0.0002, **Figure 6C**). There was a significant increase in theta power across the right centroparietal and occipital brain regions (CCP5h, FC1, C3, CP1, Pz, Oz, O2, P8, CP2, Cz, FC6, FC2, F4, FC3, C1, CP3, P1, POz, PO4, P6, TP8, FC4, F2) post-sham VR-tACS (p = 0.017, **Figure 6D**). There was no significant change in theta power when comparing stimulation conditions (see supplementary materials).

#### 3.2.2 Resting State EEG ROI Analysis

##### 3.2.2.1 Pre vs post VR-tACS

Due to a violation in the assumption of normality, Wilcoxon signed rank tests were performed to investigate the change in theta power at rest post VR-tACS compared to pre (**Figure 7**). There was a significant increase in theta power at rest at the ROI with eyes open post VR and active tACS (*z* = –3.841, *p* = <0.001) and sham (*z* = – 5.235, *p* = <0.001). There was also a significant increase in resting state theta power at rest at the ROI with resting eyes closed post VR and active tACS (*z* = –2.937, *p* = 0.002) and sham (*t*(20) = –3.286, *p* = 0.004 assumption of normality not violated).

**Figure 7:**
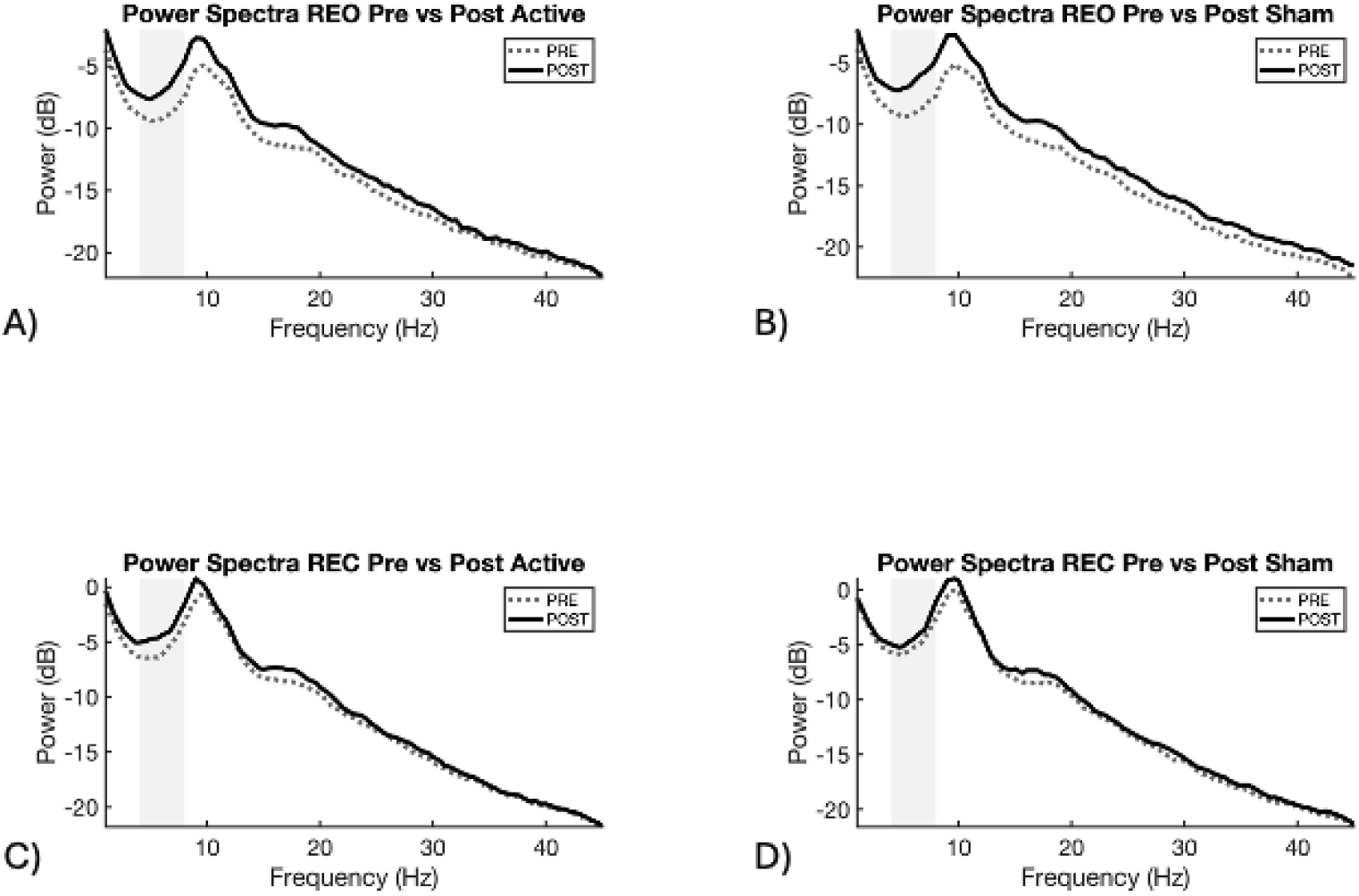
Resting state ROI power spectra plots pre compared to post VR and active or sham tACS. A and B show difference in power pre vs post VR-tACS for REO. C and D show difference in power pre vs post VR-tACS for REC. The grey bar indicates theta band activity (4-8Hz).

##### 3.2.2.2 Active vs Sham

There was a significant increase in theta power at rest with eyes closed at the rTPJ (*t* = –2.216, *p* = 0.038) in the active compared to sham session (see supplementary materials). However, normality was violated and was no longer significant when using the Wilcoxon signed-rank test (*z* = –1.234, *p* = 0.229). Bayesian analysis was conducted as a follow up which showed a potential weak support of H1 (BF_10_ = 1.681). There was no significant difference in resting eyes open between active and sham sessions (*z* = 0.678, *p* = 0.517, BF_10_ = 0.239).

#### 3.2.3 Event-Related Potentials

##### 3.2.3.1 TP450

There were no significant differences for ROI or cluster-based statistics. See supplementary materials for additional information.

##### 3.2.3.2 LPC

There were no significant differences for ROI or cluster-based statistics. See supplementary materials for additional information.

### 3.3 Correlation

Exploratory correlations were conducted to investigate the relationship between the significant findings in resting state theta activity and task performance. Due to the presence of two outliers in the data set (values 2SD from the mean), the correlations were run first with the outliers included and then with them excluded both are reported here. With the outliers retained there was a significant moderate negative correlation between change in resting eyes open theta power and change in ToM task response time for VR+tACS (r = –0.506, *p* = 0.019, BF_10_ = 3.524. **Figure 8**). With the outliers removed, the correlation was no longer significant (r = –0.244, *p* = 0.30, BF_10_ =. 0.458).

**Figure 8:**
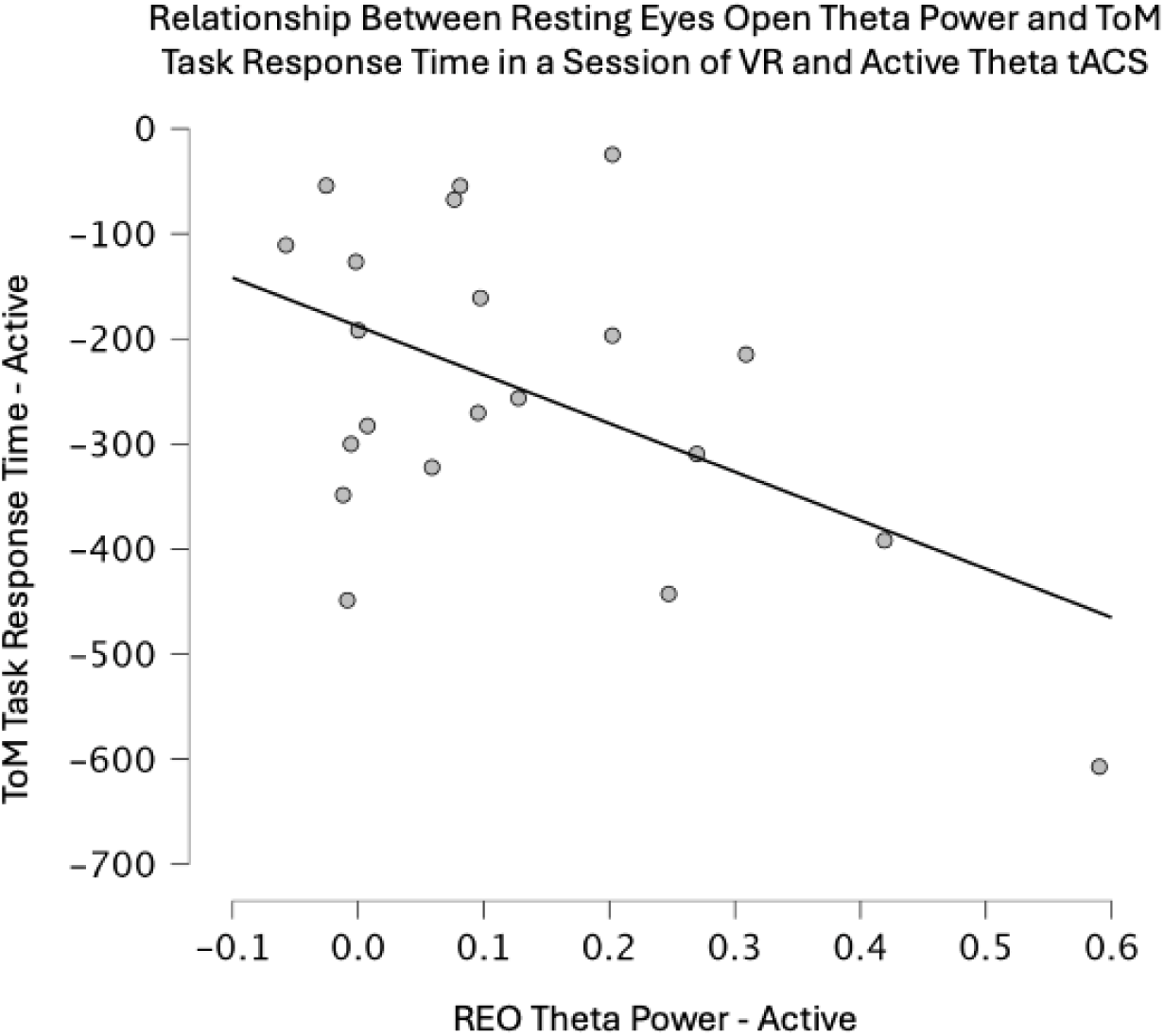
Relationship between difference in resting eyes open theta power (pre vs post) VR and active theta tACS and difference in ToM task response time (pre vs post) VR and active theta tACS.

### 3.4 tACS Blinding and Intensity

Participants failed to guess at a rate better than chance whether they were receiving sham tACS (χ^2^ (1, 21) = 1.190, *p* = 0.275), expressing a moderate degree of confidence in their guess (M = 5.667, SD = 3.246). Participants were more likely to correctly guess when they were receiving active tACS (χ^2^ (1, 21) = 10.714, *p* = 0.001), again, with a moderate degree of confidence (M = 6.000, SD = 2.702). A Wilcoxon signed rank test (used due to violation of assumption of normality) showed no significant difference in how participants perceived stimulation intensity between active and sham conditions (*z* = –1.501, *p* = 0.133).

### 3.5 VR Sickness and Immersion

Participants reported experiencing significantly less VR sickness when receiving active tACS compared to sham (Wilcoxon signed rank test, *z* = 2.10, *p* = 0.037). They also reported significantly lower total VR sickness questionnaire (VRSQ) scores during active stimulation (*t*(20) = 2.606, *p* = 0.017). There was no significant difference in oculomotor side effects (Wilcoxon signed rank test, *z* = 1.376, *p* = 0.178). Nor were there significant difference in how participants experienced presence or immersion in VR between active and sham conditions on any of the PQ sub-scores. These include realism (*t*(20) = –0.216, *p* = 0.831), possibility to act (*t*(20) = –0.686, *p* = 0.503), quality of interface (Wilcoxon signed rank, *z* = 1.041, *p* = 0.305), possibility to examine (Wilcoxon signed rank, *z* = 0.753, *p* = 0.462), self-evaluation (*t*(20) = 0.384, *p* = 0.705) and sound (*t*(20) = –1.521, *p* = 0.144). See supplementary materials for further information.

## 4 Discussion

This was the first study to investigate the effects of VR social cognition training with concurrent theta tACS on behavioural and neurophysiological outcomes. We found a significant improvement in ToM task accuracy following VR+tACS, with no improvement following VR+Sham. NToM task accuracy significantly improved following both active and sham VR-tACS. While both ToM and NToM response time also significantly improved following active and sham sessions. There were no differences *between* active and sham VR-tACS for any of the behavioural measures.

Resting state theta power significantly increased following both active and sham VR-tACS conditions, again with no significant difference *between* the stimulation conditions. Finally, results suggested the presence of a potentially underpowered relationship between change in theta power and improvement in ToM task response time in the active stimulation condition only. Self-report measures of VR and tACS additionally provided support for the feasibility, tolerability, and practicability of implementing a combined VR-tACS protocol for social cognition.

### 4.1 ToM and NToM Task Performance

There was an improvement in NToM accuracy irrespective of whether participants received active or sham stimulation. Reaction time also improved for both ToM and NToM tasks regardless of stimulation condition. These findings suggest either a practice effect or an effect of VR on task performance more broadly, or perhaps more likely a combination of the two. In a healthy sample, the expectation is that there are no deficits in learning ability and therefore practice effects are expected (Goldberg, Harvey, Wesnes, Snyder, & Schneider, 2015). It is also necessary to consider the potential for ceiling effects in a healthy control sample. There is evidence to suggest that ToM measures show ceiling effects in healthy samples (Yeung, Apperly, & Devine, 2024) which may, in this case, have limited the improvements seen over sham. Thus, there is likely to be more capacity to improve performance where there are deficits, such as in illness states. However, only VR+tACS, and not VR+Sham, resulted in significant improvements in ToM task response accuracy. This suggests tACS to the rTPJ may have successfully engaged neural processes involved in ToM, contributing to a more targeted response to the VR training. VR social cognition training and tACS on their own have both been shown to improve ToM (Arts et al., 2025; Christian et al., 2023; Pérez-Ferrara et al., 2024). While research has not yet been conducted to specifically investigate the combination of these technologies for social cognition, research in other fields such as stroke rehabilitation and phobias, have found the concurrent use of VR and NIBS to be more effective than either on their own (Cassani et al., 2020). Thus, given the novelty of this finding, further, larger scale research would need to be conducted to determine if this is a robust effect.

### 4.2 Resting State EEG

Resting state EEG cluster-based analysis as well as ROI analysis showed a significant increase in theta power post-VR-tACS compared to pre-for eyes open and eyes closed in both active and sham conditions. Because the VR program was the same in both conditions (i.e., there was no VR control condition) and it was built for participants to engage in ToM and social cognitive thought processes, this may have resulted in a change in brain activity elicited by VR alone. VR cognitive training research incorporating neurophysiological measures has consistently shown that VR alone can modulate brain activity. Casella et al. (2024) conducted a study using immersive VR reaction training to improve cognitive motor responses. Healthy participants completed one, 30-minute session per week for five weeks compared to a control group that had no motor reaction training. ERP amplitude measures after receiving training showed that anticipatory motor readiness in premotor brain areas had significantly increased compared to controls. Participants with autism spectrum disorder (ASD) who underwent 10 hrs of VR social cognition training across five weeks, showed increases in ERP peak amplitude in prefrontal and centro-parietal brain areas associated with ToM when completing fMRI tasks post-training (Yang et al., 2018). Other VR exposure therapy studies using fNIRS to measure brain changes, have shown that exposure sessions changed oxygenation in the prefrontal cortex during training and adjusted activity over time (Landowska, Roberts, Eachus, & Barrett, 2018). VR cognitive training alone has also been shown to increase frontoparietal connectivity in the theta activity band to improve attention and cognition in participants with ASD (De Luca et al., 2021). Thus, providing support for modulation of brain activity with VR alone.

The increase in theta power seen in the current study was widespread across the whole brain. There are several possible explanations for this. Firstly, social cognitive processes, particularly ToM, are complex skillsets that require the synchronous activation of multiple brain areas and networks to be effectively executed (Gainsford et al., 2020; Mossad, Vandewouw, Villa, Pang, & Taylor, 2022). Indeed, the rTPJ, our stimulation target, is considered a hub of ToM processes (Seymour et al., 2018; Wang et al., 2016) a key contributor to widespread neural network communication such as with the default mode network (DMN). The DMN is commonly associated with ToM and social cognitive processes and spans frontal (medial prefrontal cortex; MPFC) and parietal (precuneus; PC, and posterior cingulate cortex; PCC) regions (Menon, 2011; Van Overwalle, 2009). Research investigating the neural processes of ToM suggests initial attention, identification and processing of stimuli occurs in frontal regions such as the dorsolateral prefrontal cortex (DLPFC) and MPFC (Jiang, Wang, Li, & Li, 2016), then moves to central, centroparietal (PC, PCC) and temporoparietal regions (rTPJ) for engagement of specific ToM processes such as false belief reasoning and attribution of intentions (Jiang et al., 2016; McCleery, Surtees, Graham, Richards, & Apperly, 2011; Meinhardt, Sodian, Thoermer, Döhnel, & Sommer, 2011; Peng et al., 2018; Vistoli et al., 2015). Finally, frontal regions such as the MPFC activate to move between self and other interpretations (Jiang et al., 2016; McCleery et al., 2011; Meinhardt et al., 2011; Peng et al., 2018; Vistoli et al., 2015).

VR training also may be activating broad neural resources. The VR task requires participants to walk around the virtual environment, calculate how much to pay for a meal, as well as consider what virtual avatars may be thinking or feeling. Thus, requiring the engagement of cognitive, social cognitive and motor resources. To allow this, other networks such as the salience network may activate to attend to, and process social stimuli (Menon, 2011) and the central executive network may activate to engage in higher order cognitive information processing (Menon & Uddin, 2010). Other areas associated with memory and cognition such as the DLPFC (Barbey, Koenigs, & Grafman, 2013) are also likely activated as well as premotor and motor areas (Casella et al., 2024). Thus, the widespread activation seen at rest, may be a demonstration of this complex interplay involving both cognitive and social cognitive neural resources. While one significant advantage of VR is that it allows replication of real-world scenarios and improved ecological validity, it’s important to consider when investigating neurophysiological change, that a more naturalistic setting can increase the number of potential confounds. Striking a balance between these two factors will be important for ongoing research.

Finally, the relationship between the task performance and resting state findings were supported by the correlation of increased resting eyes open theta power after active VR-tACS and improvement in response time on ToM. However, this finding should be interpreted with caution due to the change in significance when accounting for outliers.

### 4.3 Event-Related Potentials

We did not see a significant effect of VR-tACS on ERPs during EEG task performance in either the active or sham sessions, despite ERPs following the expected time course and peaking at the expected time points (see supplementary material). The VR-tACS protocol may not have been activating specific ToM neural processes but rather, similarly to the resting state findings, may have been modulating neural activity more broadly. Thus, diffusing the potential to see ERP amplitude changes in a specific brain area and timeframe. Additionally, ToM neural processes are complex and consist of multitude skills, both explicit and implicit, that are utilised in each moment (Roheger et al., 2022). ToM research tries to identify specific ERP time frames that represent these complex processes but, because of this complexity, there is a lot of variability in how they are studied. In ToM studies, the LPC and TP450 ERPs (among others) are studied, but varied electrodes and differing time frames are used to measure these (e.g., McCleery et al., 2011; Meinhardt et al., 2011; Wang, Su, & Hong, 2021). Because of the heterogeneity in how ERPs are identified and measured, and because of how complex social cognitive neural processes are, it is difficult to pinpoint exact brain processes to measure one element of a complex whole.

### 4.4 VR and tACS Integration

Participants reported experiencing significantly lower disorientation VR sickness symptoms in the VR+tACS condition compared to VR+Sham, as well as lower overall VR sickness. There was no significant difference in experience of presence and immersion for participants in active vs sham sessions. Finally, participants experienced minimal tACS side effects.

Overall, these findings suggest that participants had few difficulties with carrying out the tasks required for the combined VR-tACS protocol. The significantly lower scores of disorientation VR side effects in the VR+tACS condition compared to VR+Sham, supports research showing a reduction of VR side effects when using a VR-tACS protocol (Li & Zanto, 2024). This could be an additional advantage of combining VR and tACS to consider for future interventions and it may be advantageous to include a measure of this in future studies. While the expectation would be that VR with active tACS would have resulted in increased alertness and presence, the null finding may indicate good tolerance of the combined protocol with no condition being “better” than the other. Additionally, reports of tACS side effects were low, VR side effects were low, and presence scores were high, regardless of statistical significance (see supplementary materials). This suggests that even with the tACS being administered at the same time as wearing the EEG cap, carrying the EEG cords on their back, navigating around the physical space and engaging in the virtual task, the participants were able to feel like they were present in the virtual environment and successfully complete the session. Taken together, these self-reported measures of experiences within VR and tACS, demonstrate a positive outlook for the participant-perceived experience and for the feasibility of combining these technologies for future interventions.

## 5. Limitations and Future Directions

While there was an effect of VR social cognitive training on both behavioural and neurophysiological measures, the addition of tACS, while differentially improving ToM accuracy, did not result in significant gains over and above VR. It may be that more sessions are required to have a more robust effect and future research trialling a multiple session protocol will be useful. Future research would also benefit from increased sample numbers. Some of the findings may have resulted from the study being under powered such as the lack of ERP findings, and correlation findings.

However, as a proof-of-concept, the study provides a strong base for the feasibility and further investigation of this type of VR-NIBS protocol for social cognition in larger sampled clinical trials with multiple sessions. Alongside this, improving the ToM tasks to measure changes in brain activity and task performance by including more trials could potentially make them more sensitive to neural activation. Additionally, given the results of this study suggesting an effect of VR, as well as evidence that VR alone can improve or modulate brain activity and cognition, the inclusion of a “true sham” condition with both control VR and sham tACS would help to provide more concrete evidence of the efficacy of concurrent VR-tACS. tACS blinding was only partially effective in this study with participants able to accurately determine if they were receiving active tACS better than chance. However, there was no significant difference in reported stimulation intensity. Finally, as VR-NIBS protocols become more commonly used as interventions, it would be helpful to develop specific side-effects and user experience questionnaires that account for the unique experiences of these combined protocols. These measures could assist in improving efficacy of these interventions by creating a more user-friendly, comfortable and immersive experience.

## 6. Conclusion

The current study provides evidence for the feasibility and practicability of combining VR, tACS and EEG cohesively. This research therefore provides a foundation to continue investigating the efficacy of VR social cognition training and tACS in larger scale clinical trials in healthy samples as well as in clinical populations in multi-session protocols.

## Data Availability

All data produced in the present study are available upon reasonable request to the authors

## Acknowledgements

We gratefully acknowledge the time and involvement from the participants. This research would not have been possible without them.

