## Supplementary Data for "Combined virtual reality social cognition training and theta transcranial alternating current stimulation: A proof-of-concept study in healthy controls"

1. Methods

1.1 Study Design

Research was conducted across two sites. All equipment used at both sites was the same and no participants were tested across sites. All participants completed their full three sessions at a single site.

1.2 EEG Montage

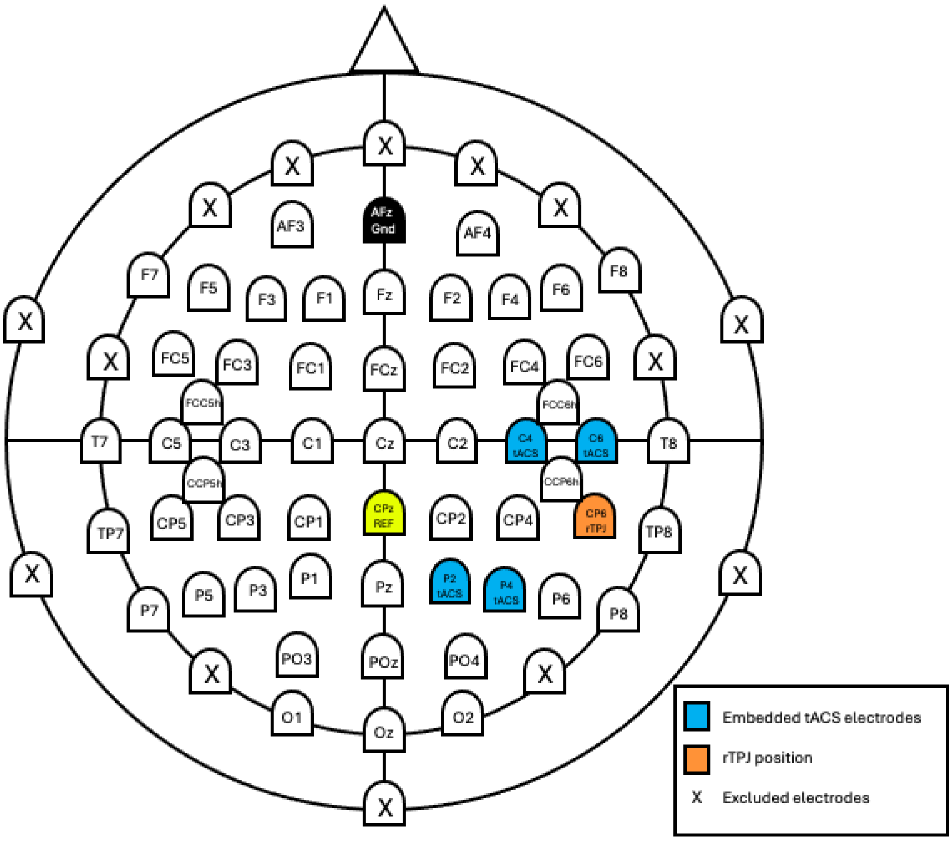


**Figure S1:** *EEG montage with embedded tACS electrodes (blue) and rTPJ highlighted (orange).* The "X's" indicate where electrodes were removed for the comfort of the VR headset (frontally) and to improve efficiency with EEG set up.

2. Analysis

2.1 Resting State Analysis Epochs

**Table S1**: *Mean number of epochs in resting state data analysis*

| | | Active | Sham |
|---|---|---|---|
| Condition | Time Point | M(SD) | M(SD) |
| REO | Pre | 56.71(2.80) | 56.10(2.76) |
| | Post | 55.57(2.44) | 55.76(3.58) |
| REC | Pre | 54.86(6.54) | 53.29(8.47) |
| | Post | 53.67(7.59) | 51.91(9.62) |

REO = resting eyes open, REC = resting eyes closed

2.2 ERP Analysis Epochs

**Table S2**: *Mean number of epochs in behavioural task data*

| | | Active | Sham |
|---|---|---|---|
| Condition | Time Point | M(SD) | M(SD) |
| ToM | Pre | 23.67(0.66) | 23.19(1.12) |
| | Post | 23.48(0.93) | 23.86(0.48) |
| NToM | Pre | 47.24(1.14) | 47.38(0.74) |
| | Post | 47.10(1.00) | 47.24(0.77) |

ToM = theory of mind task, NToM = non-theory of mind task

3. Results

3.1 Resting State Cluster Statistics

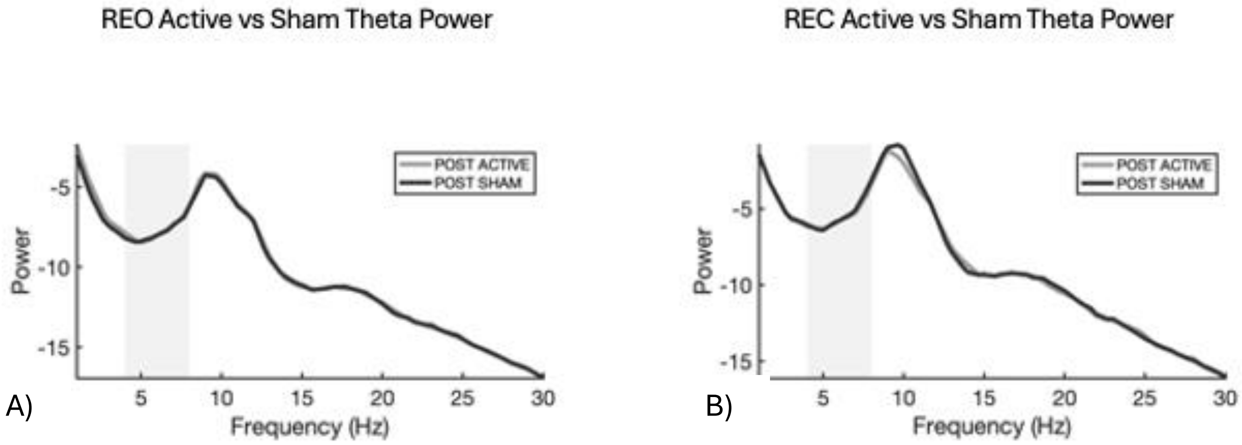


**Figure S2:** *The plots compare post VR and active tACS with post VR and sham tACS across all electrodes. The grey bar on the spectral power plots highlights the theta activity band (4-8Hz).*

3.2 Resting State ROI Analysis

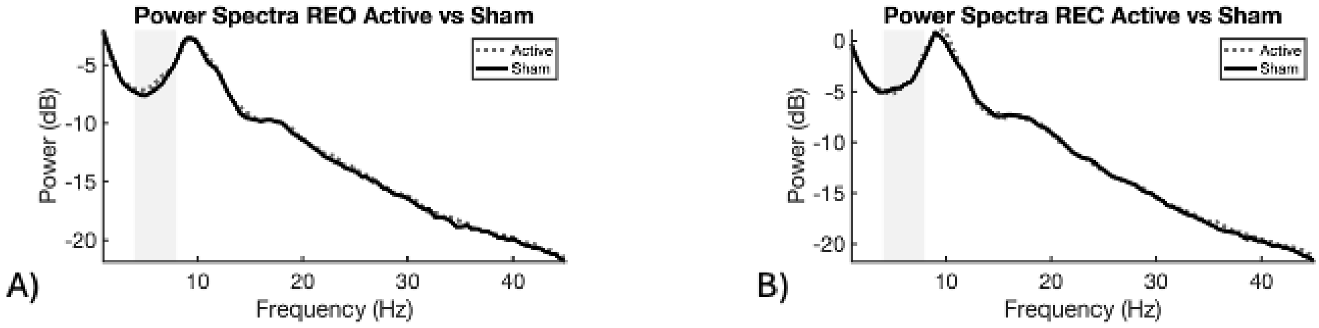


**Figure S3**: *Resting state ROI power spectra plots. A and B show difference in theta power for REO and REC in active vs sham conditions. The grey bar indicates theta band activity (4-8Hz).*

3.3 Event-related Potentials

3.3.1 TP450

There were no significant differences in theta power for the TP450 ERP at the CP6 ROI during the ToM task when comparing post VR and active tACS with pre ($t(20) = 0.167$, $p = 0.869$) or sham ($t(20) = 1.327$, $p = 0.199$). There were no significant differences in theta power for the TP450 ERP at the CP6 ROI during the NToM task when comparing post VR and active tACS with pre ($t(20) = 0.733$, $p = 0.472$) or sham ($t(20) = -0.011$, $p = 0.992$). There were no significant differences in theta power between VR and active and sham tACS sessions during the ToM task ($t(20) = -0.806$, $p = 0.430$) or the NToM task ($t(20) = 0.735$, $p = 0.471$).

There were no significant differences in theta power for the TP450 ERP at the rTPJ ROI (CP4, CP6, TP8, P6, P8) during the ToM task post VR and active tACS ($t(20) = 0.002$, $p = 0.998$) or sham ($t(20) = 0.839$, $p = 0.411$). There were no significant differences in theta power for the TP450 ERP at the rTPJ ROI (CP4, CP6, TP8, P6, P8) during the NToM task post VR and active tACS ($t(20) = 0.819$, $p = 0.423$) or sham ($t(20) = -0.297$, $p = 0.770$). There were no significant differences in theta power between VR and active and sham tACS sessions during the ToM task ($t(20) = -0.582$, $p = 0.567$) or the NToM task ($t(20) = 0.253$, $p = 0.803$).

There were no significant changes in theta power during the TP450 ERP in cluster-based analyses.

### 3.3.2 LPC

There were no significant differences in theta power for the LPC ERP at the Pz ROI during the ToM task when comparing post VR and active tACS with pre ($t(20) = -0.121$, $p = 0.905$) or sham ($t(20) = 1.783$, $p = 0.090$). There were no significant differences in theta power for the LPC ERP at the Pz ROI during the NToM task when comparing post VR and active tACS with pre (Wilcoxon signed rank, $z = 0.167$, $p = 0.191$) or sham ($t(20) = -0.292$, $p = 0.774$). There were no significant differences in theta power between VR and active and sham tACS sessions during the ToM task ($t(20) = -1.233$, $p = 0.232$) or the NToM task (Wilcoxon signed rank, $z = 0.713$, $p = 0.495$).

There were also no significant differences in theta power for the LPC ERP at the Pz, Cz ROI during the ToM task post VR and active tACS ($t(20) = -0.733$, $p = 0.472$) or sham ($t(20) = 1.585$, $p = 0.129$). There were also no significant differences in theta power for the LPC ERP at the Pz, Cz ROI during the NToM task post VR and active tACS (Wilcoxon signed rank, $z= 1.025$, $p = 0.320$) or sham ($t(20) = -0.227$, $p = 0.823$). There were no significant differences in theta power between VR and active and sham tACS sessions during the ToM task ($t(20) = -1.693$, $p = 0.106$) or the NToM task (Wilcoxon signed rank, $z = 0.574$, $p = 0.585$).

There were no significant changes in theta power during the LPC ERP in cluster-based analyses.

### 3.3.3 Cluster Statistics ERPs

There were no significant clusters in any of the conducted analyses pre vs post VR with active or sham tACS as well as between active and sham sessions for any of the investigated ERPs. **Figure 2** below provides an illustration of the ERPs being activated but without significant differences in amplitude between groups.

ToM TP450 ERPs

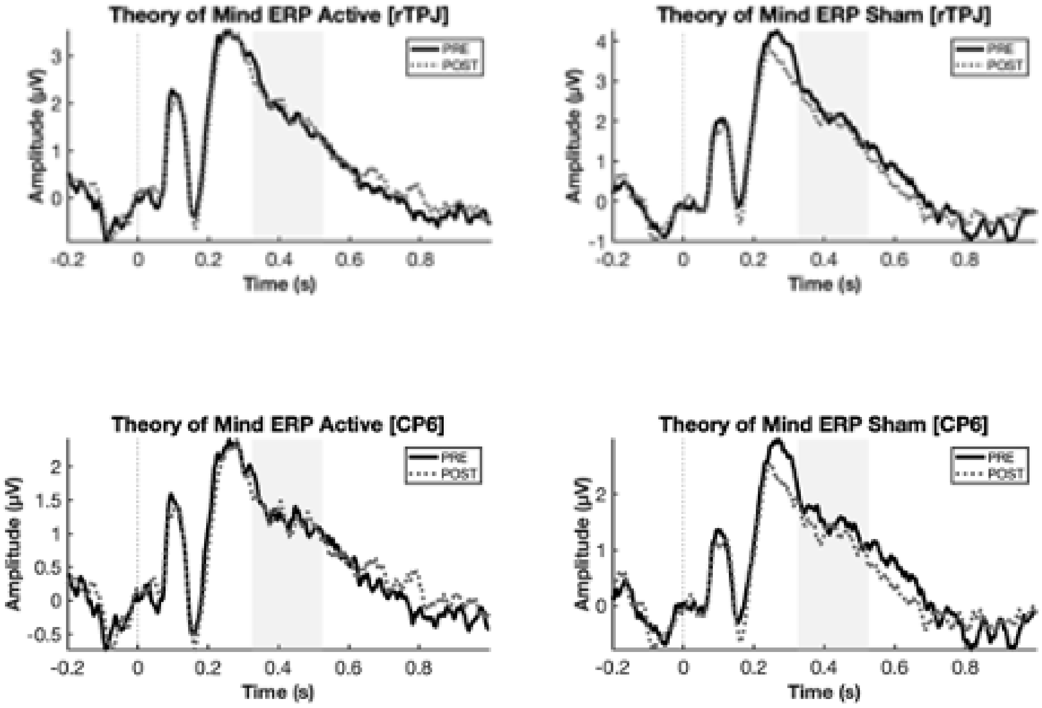


A)

NToM TP450 ERPs

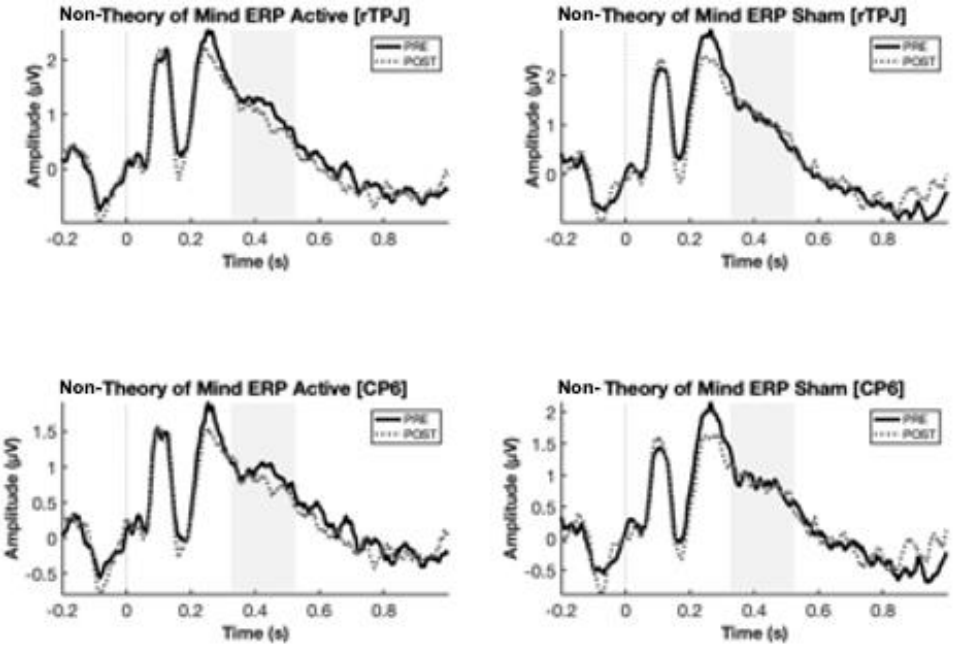


B)

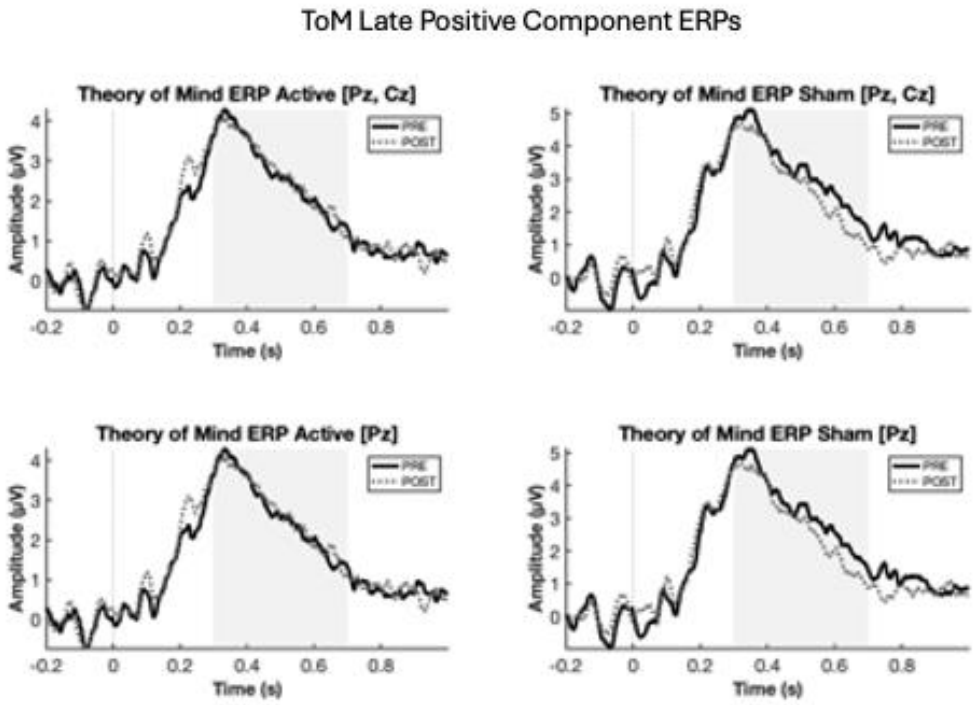


C)

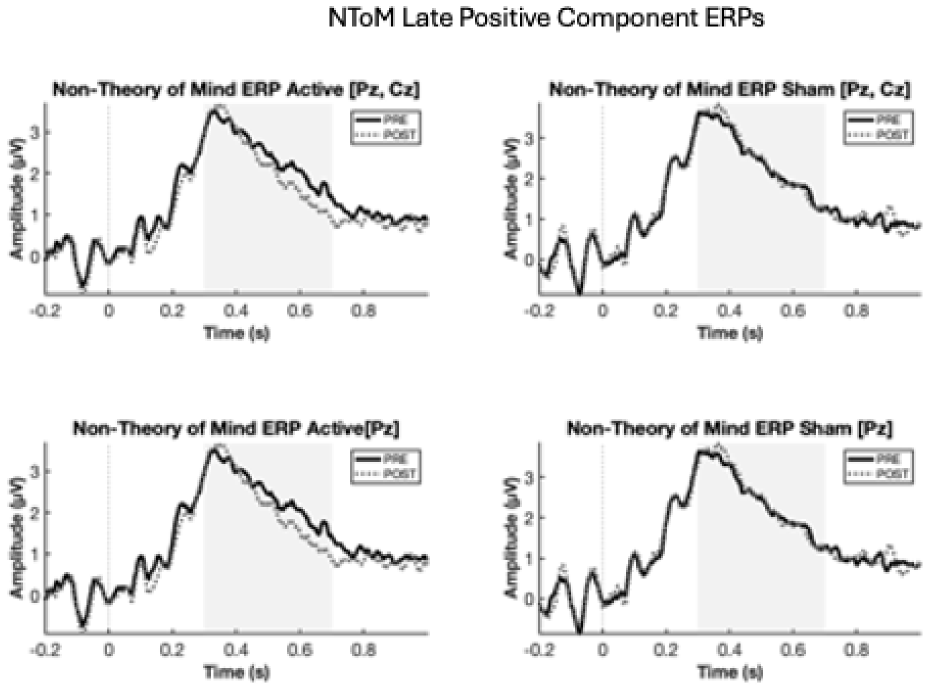


D)

**Figure S4**: *ERPs pre vs post VR and active and sham tACS*. TP450 power spectra plots (a, b) highlight the TP450 time frame (325-525ms). Late positive component (LPC) plots (c, d) highlight the LPC time frame (300-700ms). All plots investigate theta power (4-8Hz) mean amplitude at the designated electrodes. There was no significant difference for any of the ERPs measured.

3.4 VR Side Effects and Presence

**Table S3**: *Descriptive statistics of VR measures*

| | | Active | | Sham | |
|---|---|---|---|---|---|
| Measure | Sub-score | M(SD) | Range | M(SD) | Range |
| VR Sickness Questionnaire | Oculomotor | 4.365 (7.274) | 0-25 | 7.143(8.853) | 0-25 |
| | Disorientation | 1.905 (4.781) | 0-20 | 4.762(6.714) | 0-20 |
| | Total | 3.135 (5.068) | 0-18.33 | 5.952(6.386) | 0-19.17 |
| Presence Questionnaire | Realism | 37.238 (5.603) | 27-45 | 37.048 (6.012) | 22-47 |
| | Possibility to Act | 23.714 (3.196) | 18-28 | 23.286 (2.452) | 19-28 |
| | Quality of Interface | 15.571 (2.749) | 11-21 | 16.048 (3.383) | 6-21 |
| | Possibility to Examine | 17.333 (2.798) | 12-21 | 17.714 (2.194) | 14-21 |
| | Self-evaluation | 12.952 (1.024) | 11-14 | 13.048 (1.071) | 11-14 |
| | Sounds | 17.762 (3.727) | 6-21 | 17.095 (3.520) | 8-21 |
